# Cerebral Oxygenation Stability In Extremely Preterm Infants: A Randomized Clinical Trial

**DOI:** 10.1101/2025.10.21.25338440

**Authors:** Pranav R Jani, Traci-Anne Goyen, Kiran Kumar Balegar, Rajesh Maheshwari, Maria Saito-Benz, Tim Schindler, James Moore, Manelle Merhi, Melinda Cruz, Yang Song, Hayley McDonagh, Melissa Luig, Mark Tracy, Daphne D’Cruz, Aldo Perdomo, Stephanie Morakeas, Vishnu Dasireddy, Mihaela Culcer, Vijay Shingde, Karen Bennington, Joanna Michalowski, Andreja Fucek, Jennifer Querim, Sean Stevens, James Santanelli, James Elhindi, Brian Gloss, Robert Halliday, Dharmesh Shah, Himanshu Popat

**Affiliations:** Department of Neonatology, Westmead Hospital, Westmead, Australia; Faculty of Medicine and Health, The University of Sydney, Sydney, Australia; Department of Neonatology, Nepean Hospital, Nepean Blue Mountains Local Health District, Kingswood, Australia; Sydney Medical School Nepean, The University of Sydney, Sydney, New South Wales, Australia; Neonatal Intensive Care Unit, Wellington Regional Hospital, Wellington, New Zealand; Department of Paediatrics and Child Health, University of Otago, Wellington, New Zealand; Department of Newborn Care, The Royal Hospital for Women, Sydney, Australia; School of Clinical Medicine, University of New South Wales, Sydney, Australia; Connecticut Children’s, Division of Neonatal-Perinatal Medicine, Connecticut Children’s Medical Center, Hartford, Connecticut, USA; UCONN School of Medicine Farmington, Farmington, Connecticut, USA; Kennards Hire Group, Australia; NHMRC Clinical Trial Centre, University of Sydney, Camperdown, Australia; NICU Lived Network, Australia; Lived Experience Advisory Network, Perinatal Society of Australia and New Zealand; Research and Education Network, Westmead Hospital, Westmead, Australia; Faculty of Engineering and Information Technologies, BMET Institute, The University of Sydney, Sydney, New South Wales, Australia; Westmead Research Hub, Westmead Institute for Medical Research, Sydney, New South Wales, Australia; Grace Centre for Newborn Intensive Care, The Childre’s Hospital for Westmead, Westmead, Australia

## Abstract

**Importance:** Preterm infants are at high-risk of developing brain injury. Near-infrared spectroscopy (NIRS) offers the ability to measure cerebral oxygenation, potentially reducing brain injury. What remains unknown is the impact of using a standardized treatment guideline combined with a single NIRS device manufacturer and neonatal sensor on cerebral oxygenation, which has not been previously examined.

**Objective:** To determine whether cerebral NIRS monitoring with a dedicated treatment guideline improves cerebral oxygenation stability.

**Design:** This was a single-blinded, two-arm randomized controlled trial conducted from October 2021 to July 2024.

**Setting:** Five tertiary neonatal intensive care units across Australia, New Zealand and the United States.

**Participants:** Infants born <29 weeks’ gestation and <6 hours of age underwent 1:1 random allocation, stratified by gestational age (<26 weeks’ and ≥26 weeks’) and study site.

**Intervention:** The intervention group received **c**erebral NIRS monitoring and dedicated guideline-based treatment when the cerebral oxygenation was outside the range of 65%–90%. The control group had blinded cerebral NIRS monitoring and treatment guided by standard clinical monitoring.

**Main Outcome(s) and Measure(s):** The burden of cerebral hypoxia and hyperoxia during the first 5 days after birth expressed as percent hours was the primary outcome. Key secondary outcomes were mortality, morbidities before discharge, and NIRS-related skin injury.

**Results:** Of the 149 screened infants, 100 were included in the final analysis. The median gestational age was 27 weeks’ (inter quartile range [IQR 25–28]) and the median birth weight was 883 grams (IQR 709–1079). The intervention group (n=50) had a significantly lower median burden of hypoxia and hyperoxia of 5.7% hours (IQR 2.8–15) compared to 39.6% hours (IQR 6.5–82.3) in the standard care group (n=50), with an adjusted reduction of 42.8% hours (95% confidence interval 35.6–53.3, p=0.0002). Mortality, morbidities before discharge and safety outcomes were comparable between groups.

**Conclusions and Relevance:** Treatment guided by cerebral NIRS monitoring with a single device manufacturer and neonatal sensor, is a safe and low-risk intervention that significantly improves stability of cerebral oxygenation in extremely preterm infants. Larger multicenter trials are warranted to determine if this finding leads to improved survival without brain injury.

**Trial Registration:** The trial is registered at The Australian New Zealand Clinical Trials Registry, registration number: ACTRN12621000778886, and https://www.anzctr.org.au/Trial/Registration/TrialReview.aspx?ACTRN=12621000778886

**Key points:** *Question:* Does cerebral near infrared spectroscopy (NIRS) with a dedicated treatment guideline using NIRS device from a single manufacturer and neonatal sensor improve cerebral oxygenation stability in extremely preterm infants?

*Findings:* In this randomized clinical trial of 100 infants, the burden of cerebral hypoxia and hyperoxia was significantly lower in the intervention group (5.7% hours) compared to standard care group 39.6% hours.

*Meaning:* Treatment guided by cerebral NIRS monitoring is a safe and low-risk intervention that improves stability of cerebral oxygenation in extremely preterm infants.

## Introduction

Extremely preterm (<28 weeks’ gestation) infants have a higher risk of death, neurodevelopmental deficits and consequently, substantial societal economic impacts.^1–4^ They often experience cardiorespiratory instability during the early postnatal period presenting as episodes of systemic hypoxia and/or hyperoxia, both of which are associated with an increased risk of brain injury.^5–7^ A meta-analysis from five clinical trials comparing lower (85–89%) versus higher (91–95%) peripheral oxygen saturation target ranges in extremely preterm infants reported reduced risk of death and severe necrotizing enterocolitis, and greater risk of treated retinopathy of prematurity in infants assigned to the higher target ranges but similar rates of death or major disability at 18–24 months of corrected age.^8^ Targeting the stability of cerebral oxygenation by using near infrared spectroscopy (NIRS), compared to systemic assessment of oxygenation using pulse oximetry, offers an alternative physiological approach for reducing brain injury.^9^

The SafeBoosC-II study, a landmark trial, demonstrated stability of cerebral oxygenation in extremely preterm infants by using cerebral NIRS monitoring guided dedicated treatment.^10^ They used various manufacturers of NIRS platforms with modified alarm thresholds and adult sensors measuring lower absolute oxygenation values compared with neonatal sensors. This approach introduced greater variability in NIRS monitoring. Building on this evidence, the current trial, Near Infra-Red Spectroscopy Targeted Use to Reduce adverse outcomes in Extremely preterm infants (NIRTURE), tested the hypothesis that the burden of cerebral hypoxia and hyperoxia could be reduced by combining cerebral NIRS monitoring with a dedicated treatment guideline using a NIRS device from one manufacturer and a neonatal sensor, compared to blinded cerebral NIRS monitoring and standard care during the early postnatal period. As against the SafeBoosC-II study, this trial extended monitoring from 3 to 5 days after birth, used real-time, unmodified device alarms for detecting cerebral hypoxia and/or hyperoxia, NIRS device from a single manufacturer to reduce potential variability between devices and 65%–90% as the reference range for cerebral oxygenation. This trial aims to inform future RCT evaluating the impact of this intervention on childhood survival free of neurological deficits.

## Methods

The trial protocol, including details of the methodology, was previously published.^11^

### Consumer Involvement

Consumers with a lived experience of preterm birth were actively engaged in co-designing this trial. Their involvement included reviewing and advising on the study methodology, contributing to developing the consent process, particularly advocating for deferred consent, and participant information and consent forms. Both individual consumers and consumer organizations were consulted for the decision to allocate siblings of multiple births to the same treatment group.

### Trial Design

This is a multisite, single-blinded, two-arm RCT with 1:1 allocation, stratified by gestational age (<26 weeks’ and ≥26 weeks’) and study site.

### Changes to trial protocol

A modification to the trial protocol was implemented to address instances where participants had been randomized, but parental consent could not be obtained in the first 5 days after birth due to circumstances such as COVID-19 related isolation or maternal sickness requiring intensive care unit admission. In these cases, consent was later obtained from the family before including the participant’s data in the analysis. No changes were made to the predefined outcomes or the analyses after trial commencement.

### Trial setting

This trial was conducted across five tertiary units across Australia, New Zealand and the United States. The participating sites included Westmead Hospital, Nepean Hospital and The Royal Hospital for Women in Australia; Wellington Hospital in New Zealand; and The Connecticut Children’s Hospital in the United States. Investigators at each site had prior experience with cerebral NIRS monitoring. However, most clinical staff at these sites did not have previous experience in cerebral NIRS monitoring. Therefore, clinical staff underwent structured training and completed web-based certification before site activation and participant recruitment. Ethical approval was obtained in each participating country before trial commencement (Australia: Sydney Children’s Hospitals Network Human Research Ethics Committee: 2020/ETH02903; New Zealand: Health and Disability Ethics Committee: 21/NTB/157; United States: Institutional Review Board of the Connecticut Children’s Medical Center: IRB 21-071). The trial was conducted in accordance with the International Council for Harmonisation guidelines for Good Clinical Practice.

### Eligibility criteria

We included inborn and outborn infants (singleton or twin births) born <29 weeks’ gestation who were <6 hours of age. Exclusion criteria were antenatal or postnatal diagnosis of a congenital anomaly requiring major surgery, genetic disorders associated with neurological impairment and multiple births beyond twins.

### Intervention and comparator

#### Interventions

After enrolment, infants were randomly allocated to either the standard care group or the intervention group. For both groups, cerebral oxygenation monitoring was commenced by placing a neonatal NIRS sensor on the fronto-temporal region of the infant’s head on either side. Cerebral NIRS data were continuously recorded using the SenSmart Model X-100 Universal Oximetry System (NONIN Medical Inc) for 5 days from birth (120 hours). In the intervention group, real-time cerebral oxygenation readings were visible to the clinical staff, who managed infants according to a predefined treatment guideline when cerebral oxygenation values were outside the target range of 65%–90%. In the standard care group, the NIRS monitor screen was covered with an opaque cover to prevent clinical staff from acting on these values. Infants in the standard care group received care according to the participating sites’ established clinical monitoring and treatment protocols. There were no restrictions on any other aspects of concomitant care.

### Intervention thresholds

A pragmatic, consensus-based approach was used to define the reference range for cerebral oxygenation. The lower threshold of 65% was chosen based on the threshold used in the SafeBoosC-II study and adjusted to account for higher absolute oxygenation values observed with neonatal sensors. The upper threshold of 90% instead of 85%, used in the SafeBoosC-II study, was chosen from local, unpublished data from Westmead Hospital. In this cohort, a cerebral oxygenation value of 87% corresponded to the 75^th^ centile. The sensor site was inspected every 4 hours to ensure correct sensor placement and skin integrity.

A clinical treatment guideline for cerebral hypoxia was activated when cerebral oxygenation (CrSO_2_) was <65% and a clinical treatment guideline for cerebral hyperoxia was activated when CrSO_2_ was >90%. Further details on the treatment guideline are available in Jani et al.^11^

### Outcomes

Primary outcome: The burden of cerebral hypoxia and hyperoxia during the first 5 days after birth, expressed as percent hours. An hour event with a mean CrSO_2_ of 55% would equate to 10% hours of hypoxia (10% below the lower threshold of 65% multiplied by 1 hour). Only deviations lasting longer than one minute were included in the calculation of primary outcome. The primary outcome for events exceeding 10 minutes is reported for comparison with other studies (eTable 1 in the supplement).^10^

Secondary outcomes before hospital discharge were mortality, brain injury on imaging (any intraventricular hemorrhage^12^ or cerebellar hemorrhage or periventricular leukomalacia), chronic lung disease–need for respiratory support or supplemental oxygen at 36 weeks’ postmenstrual age, necrotizing enterocolitis using Bell’s classification^13^ and retinopathy of prematurity (ROP).

### Harms

Skin injury (pressure or thermal) associated with the NIRS sensor within the first 5 days after birth was monitored as a safety outcome.

### Sample Size

Based on the results of the SafeBoosC-II study,^10^ we assumed that a 50% reduction in primary outcome would represent a clinically meaningful effect. This corresponded to a reduction of 0.3 in the mean of log-transformed burden of cerebral hypoxia and/or hyperoxia. Assuming a standard deviation of 0.5 in log-transformed burden,^14^ a sample size of 45 infants per group could achieve 80% power with a two-sided alpha of 0.05. Accounting for the clustering effect from twin births allocated to the same treatment and based on a regional incidence of twins of ∼ 30%, the average cluster size was estimated at 1.3 with an assumed intra-cluster correlation of 0.1. This gave a design effect of 1.03, taking the sample size to 47 per group or 94 in total. To accommodate loss to follow-up or missing data, the final sample size was set at 100 infants.

For consented infants in whom NIRS monitoring was discontinued earlier than the planned five-day after birth monitoring period, data collected until discontinuation were included in the primary outcome analysis. Study oversight was provided by an independent Data Safety and Monitoring Committee, which monitored all aspects of trial conduct 6-monthly until recruitment was complete. An interim analysis was not conducted during the trial.

### Randomization

The site investigators or clinical staff randomized participants using REDCap electronic data capture tools (University of Sydney licence), which used variable block randomization stratified by gestational age and study site. Randomization codes were generated by an independent biostatistician using permuted blocks to maintain group balance in a 1:1 allocation ratio. Twins were assigned to the same treatment group to account for clustering. Withdrawn infants were replaced in the study with new randomization. Data analysts were blinded to the treatment allocation.

### Statistics

Statistical analyses were conducted using RStudio.Version(4) at a significance level of 0.05 with a two-sided alternative hypothesis. No corrections for multiple hypotheses were performed. Participant demographic, clinical characteristics and study outcomes are presented using standard descriptive statistics: frequencies and percentages for categorical variables, mean, standard deviation and range or median, quartiles and range for continuous variables. All efficacy analyses were performed according to randomized treatment. We used a lognormal transformation to ensure normality for the primary outcome and used a linear mixed effects model with clustering by hospital for comparison between groups. Sub-group analysis for gestational age stratum was performed. Other study outcomes were compared using Linear mixed effects models with normal, lognormal or binary outcome distribution. Adjusted models were used to explore predictors of outcomes. Variables that differed significantly between the two groups, and were plausibly contributory to the outcome, were considered for adjustment. A fixed adjustment for gestational age category was included as an a priori specified variable. Models additionally including clustering for twins were examined but did not better fit the data as per a likelihood ratio test.

## Results

Between October 2021 and July 2024, a total of 149 infants were screened for eligibility. Of these, 104 were randomized to either the intervention group (n=53) or the standard care group (n=51) (Figure 1). Following randomization, four infants were excluded: two died within the first 5 days before consent could be obtained (unrelated to the intervention), one was diagnosed postnatally with a congenital anomaly requiring surgery, and one parent withdrew consent. A total of 100 infants (50 in each group) received the allocated intervention and were included in the primary outcome analysis. One infant in each group died within the first 5 days after birth and they were included in the final analyses. These deaths were not related to the intervention. Except for these two infants, all remaining infants received protocol defined cerebral NIRS monitoring for the first 5 days after birth.

**Figure 1:**
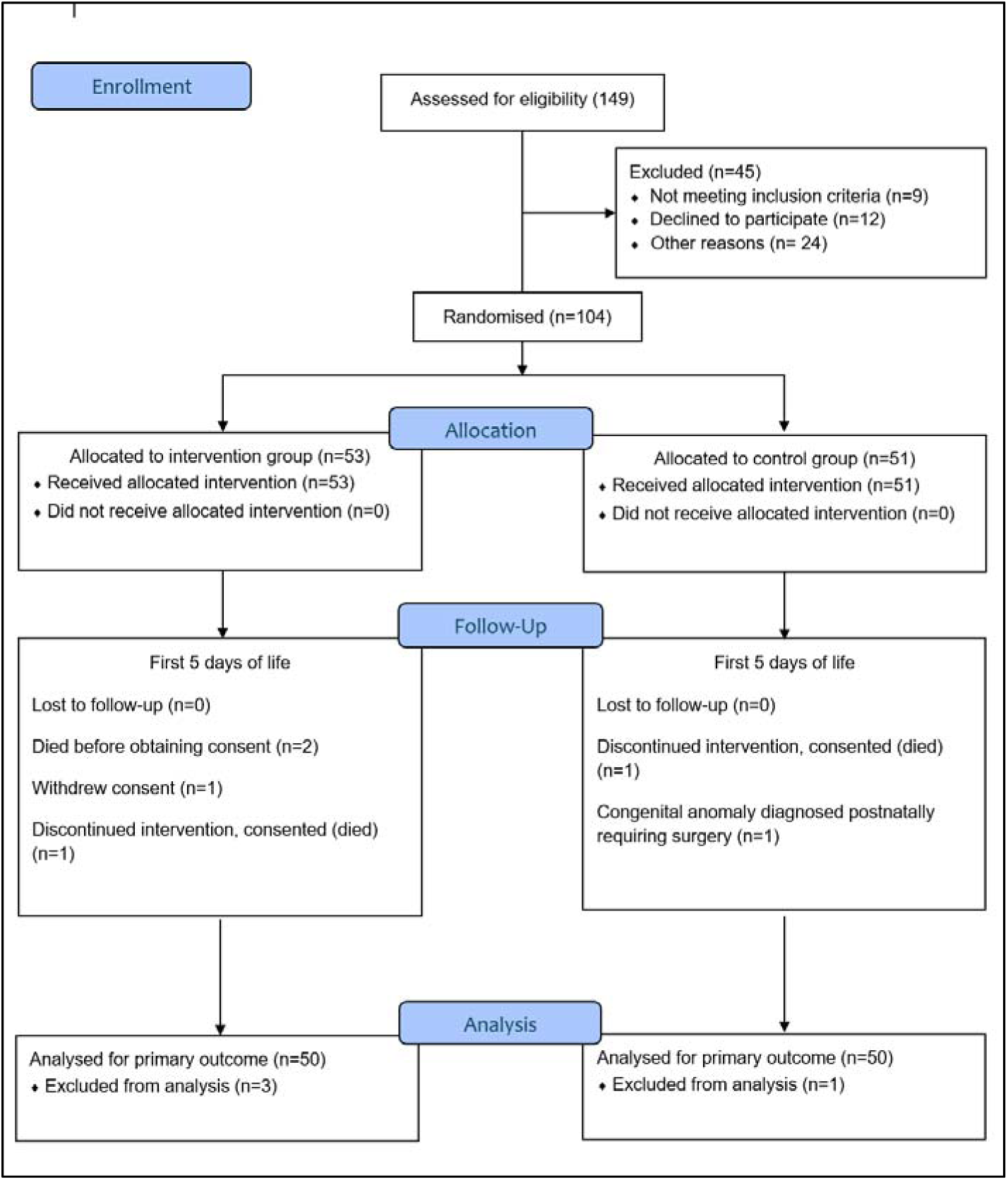
Flow of study infants as per updated CONSORT reporting guideline^19^. Other reasons: missed eligible participants, no research staff after hours for consenting Recruitment by sites: 25 at site A, 20 each at sites B and C, 32 at site D and 3 at site E

Baseline characteristics between the two groups were similar, except for a higher proportion of infants in the intervention group exposed to chorioamnionitis and of larger birth weight centiles (Table 1). These variables were adjusted for in the primary outcome analysis. The median gestational age was 27 weeks’ (interquartile range 25–28), the median birth weight was 883 g (interquartile range 709–1079) and 48 (48%) were males. No significant differences were observed between groups regarding the rates of cardiorespiratory treatment, complications, sepsis, or mortality within the first 5 days after birth (eTable 2 in the supplement).

**Table 1:**
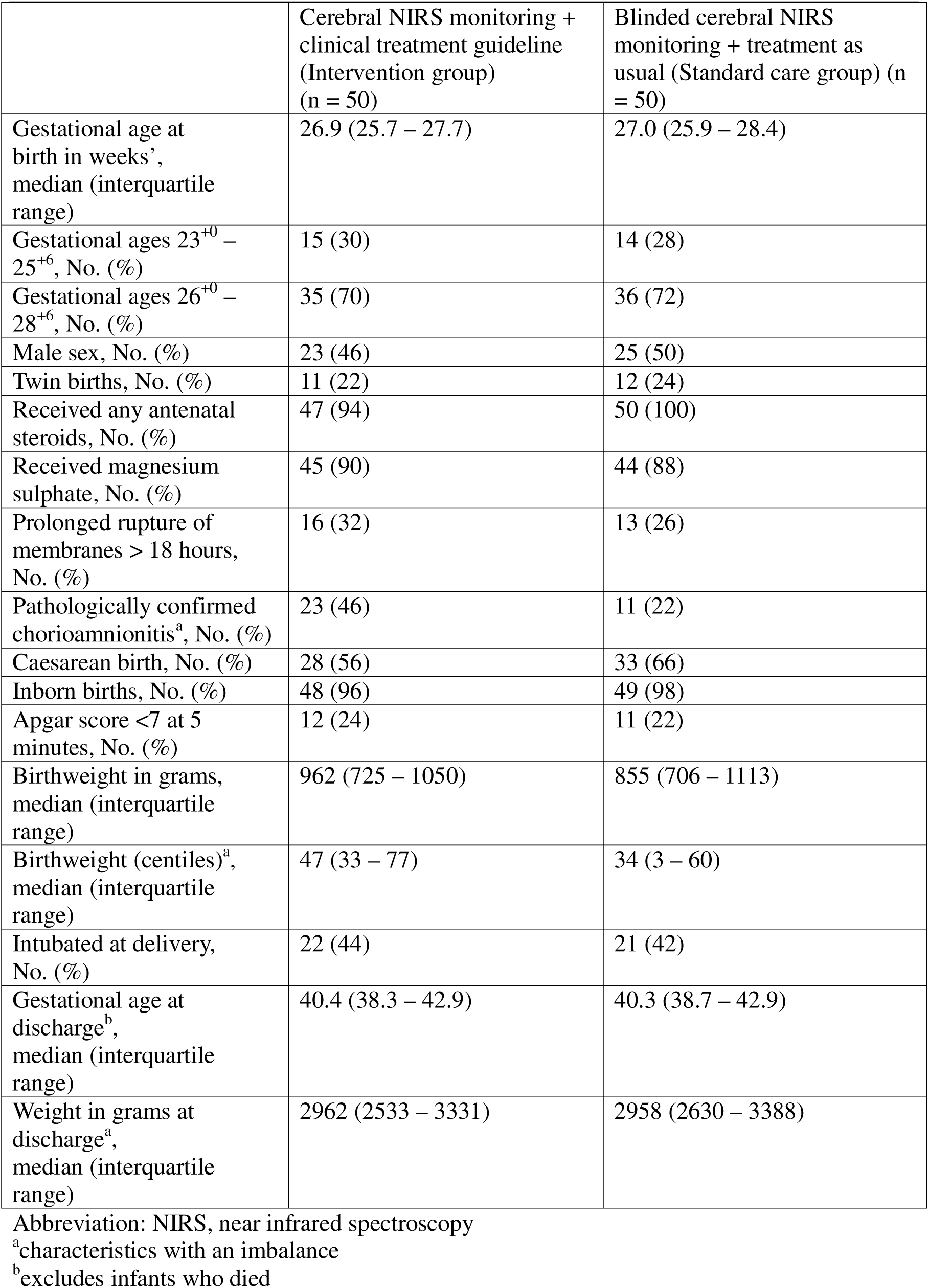
Participant characteristics.

### Primary outcome

Among the 50 infants randomized to the intervention group, the combined burden of cerebral hypoxia and hyperoxia was 5.7% hours (interquartile range [IQR] 2.8–15), compared to 39.6% hours (IQR 6.5–82.3) in 50 infants randomized to the standard care group. The adjusted relative change in the primary outcome for the intervention group was 42.8% (95% confidence intervals 35.6 to 53.3, p = 0.0002) (Figure 2). The burden of cerebral hyperoxia decreased from 23.7% hours (IQR 5.8 – 60.7) in the standard care group to 3.5% hours (IQR 2.0–14.4) in the intervention group, corresponding to an adjusted relative change of 40.3% hours (95% confidence intervals [CI] 30.5 to 59.5, p = 0.01). Similarly, the burden of cerebral hypoxia was reduced from 2.5% hours (IQR 1.1–8.0) in the standard care group to 0.7% hours (IQR 0.04–1.9) in the intervention group, with an adjusted relative change of 335.9% hours (95% CI 325.7 to 1019, p = 0.0007). Detailed results stratified by gestational age are presented in Table 2.

**Figure 2:**
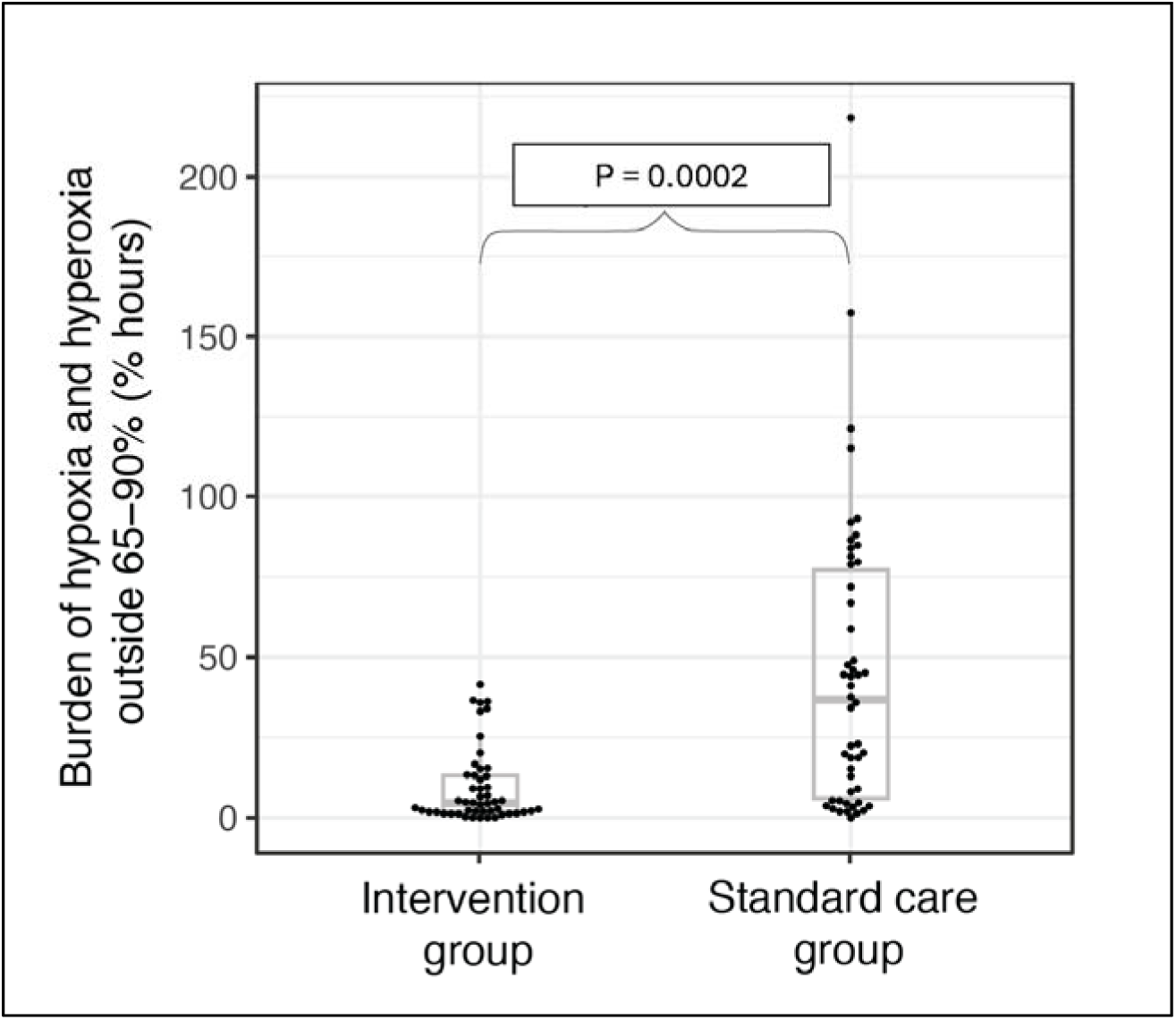
Primary outcome by treatment

**Table 2:** Primary outcome stratified by gestational age of the participants.

| Outcomes | Intervention group | Standard care group | Relative change in % (95% CI)<sup>a</sup> |
| --- | --- | --- | --- |
| **For Gestational ages 23<sup>+0</sup> – 25<sup>+6</sup>** | **n = 15** | **n = 14** |  |
| Burden of cerebral hypoxia and hyperoxia expressed as % hours, median (interquartile range) | 5.0<br>(2.9 to 13.0) | 51.6<br>(38.5 to 96.4) | 57.0<br>(47.5 to 71.0) |
| Burden of cerebral hypoxia expressed as % hours, median (interquartile range) | 1.1<br>(0.3 to 2.2) | 5.3<br>(2.6 to 18.2) | 222.6<br>(82.9 to 487.5) |
| Burden of cerebral hyperoxia expressed as % hours, median (interquartile range) | 3.3<br>(2.3 to 10.9) | 47.2<br>(23.3 to 87.3) | 61.8<br>(50.0 to 81.1) |
| **Gestational ages 26<sup>+0</sup> – 28<sup>+6</sup>** | **n = 35** | **n = 36** |  |
| Burden of cerebral hypoxia and hyperoxia expressed as % hours, median (interquartile range) | 5.9<br>(2.5 to 14.7) | 23.8<br>(5.8 to 67.0) | 32.1<br>(24.8 to 44.4) |
| Burden of cerebral hypoxia expressed as % hours, median (interquartile range) | 0.7<br>(0.04 to 1.6) | 1.8<br>(1.0 to 7.1) | 465.1<br>(134.5 to 525.8) |
| Burden of cerebral hyperoxia expressed as % hours, median (interquartile range) | 4.5<br>(1.9 to 14.1) | 13.4<br>(4.2 to 44.0) | 21.1<br>(13.9 to 40.8) |

Abbreviation: CrSO<sub>2</sub>, cerebral oxygenation; CI, confidence interval
<sup>a</sup>Model adjusted for gestation, chorioamnionitis and birthweight centile

### Secondary outcomes

No significant differences were observed between the two groups for the secondary outcomes assessed before hospital discharge (Table 3).

**Table 3:** Secondary outcomes before hospital discharge.

| Outcomes | Intervention group (n=50) | Standard care group (n=50) | Adjusted relative risk (95% CI)<sup>a</sup> |
| --- | --- | --- | --- |
| Mortality before home discharge, No. (%) | 3 (6) | 3 (6) | 1.28 (0.21 to 7.92) |
| Brain injury before hospital discharge, No. (%) | 16 (32) | 17 (34) | 0.77 (0.30 to 1.94) |
| Any intraventricular hemorrhage, No. (%) | 16 (32) | 16 (32) | 0.88 (0.35 to 2.20) |
| Severe intraventricular Hemorrhage (grades III and IV), No. (%) | 3 (6) | 2 (4) | 1.38 (0.18 to 12.94) |
| Cerebellar hemorrhage, No. (%) | 0 | 2 (4) | 1.32 (0.14 to 12.68) |
| White matter injury, No. (%) | 1 (2) | 2 (4) | 0.27<sup>b</sup> |
| Chronic lung disease at 36 weeks' postmenstrual age<sup>c</sup>, No. (%) | 27/46 (59) | 31/49 (63) | 0.91 (0.37 to 2.28) |
| Necrotizing enterocolitis ≥ stage 2, No. (%) | 3 (6) | 3 (6) | 0.84 (0.16 to 4.38) |
| Retinopathy of prematurity, No. (%) | 29 (58) | 32 (64) | 0.97 (0.90 to 1.04) |
| Surgery for retinopathy of prematurity, No. (%) | 2/29 (7) | 5/32 (16) | 0.35 (0.04 to 1.90) |
| Anti-vascular endothelial growth factor therapy for retinopathy of prematurity, No. (%) | 3/29 (10) | 6/32 (19) | 0.42 (0.06 to 2.31) |

Data is presented as number (percentage)
<sup>a</sup>Model adjusted for gestation, chorioamnionitis and birthweight centile
<sup>b</sup>Model did not converge
<sup>c</sup>Excludes infants who died before 36 weeks' post menstrual age

### Safety

No participant in the intervention group developed skin injury from the NIRS sensor. One participant in the standard care group developed a transient imprint without skin injury from the NIRS sensor. The data safety committee did not identify any additional safety issues to the infants.

## Discussion

In this trial, we demonstrated that treatment guided by cerebral NIRS monitoring, compared with treatment based on conventional clinical monitoring and blinded cerebral NIRS monitoring, significantly improved the stability of cerebral oxygenation during the first five days after birth in premature infants born <29 weeks’ gestation.

From a safety perspective, no infant in the intervention group experienced skin injury related to cerebral NIRS monitoring throughout the intervention period. In contrast, one infant in the standard care group developed a transient skin imprint from the NIRS sensor, which resolved following sensor repositioning. There were no significant differences between groups in key secondary outcomes related to major morbidities before hospital discharge. However, treatment rates for advanced ROP were non-significantly lower in the intervention group compared with the standard care group. No site used automated control of oxygen delivery during the intervention period. As the study was not powered for detecting differences in secondary outcomes such as advanced ROP requiring treatment, this finding needs exploration in future trials, including when using automated control of oxygen delivery.

The findings from the NIRTURE trial provide valuable insights into the stability of cerebral oxygenation in extremely preterm infants during the transitional phase. The SafeBoosC-II study previously demonstrated significantly improved stability of cerebral oxygenation from the intervention.^10^ In SafeBoosC-II study, cerebral hypoxia, but not cerebral hyperoxia, contributed to the primary outcome.^10^ The improvement in primary outcome in the NIRTURE trial was predominantly driven by a reduction in cerebral hyperoxia, although a significant decrease in cerebral hypoxia was also observed.

While both, the NIRTURE and SafeBoosC-II trials, demonstrated that the combination of cerebral NIRS monitoring and a dedicated treatment guideline, improved cerebral oxygenation stability with no adverse effects from the intervention. It is important to acknowledge the methodological differences between the two trials, which may influence the interpretation and generalizability of their findings. Compared to SafeBoosC-II study, the NIRTURE trial’s methods included (i) using single NIRS device manufacturer with neonatal-population specific sensor, (ii) an extended intervention period covering from 72 hours after birth to include the first five days after birth, (iii) real-time, unmodified device alarms for detecting cerebral hypoxia and/or hyperoxia (iv) eliminated device algorithm variability by using NIRS device from one manufacturer (v) using 65–90% as the reference range for cerebral oxygenation and (vi) participating site investigators, but few clinical staff, had prior experience with cerebral NIRS monitoring. The rationale for the above listed methods of the NIRTURE trial is detailed in Jani et al.^11^ While the SafeBoosC-II study demonstrated stability of cerebral oxygenation from the intervention in the first 72 hours after birth, the NIRTURE trial demonstrated further extension of this finding to the first 5 days after birth. The absence of significant differences in major morbidities before hospital discharge suggests that the intervention is safe in this vulnerable population.

The major strengths of our trial include: (1) using a commercially available, unmodified, single type of NIRS device with a neonatal-specific sensor for continuous cerebral NIRS monitoring; (2) active involvement of consumers with a lived experience of preterm birth who were integral to study from conceptualization through to development; (3) minimizing bias by blinding the data analysts to treatment allocation; (4) multisite recruitment and two consenting approaches facilitated the inclusion of infants born after hours and under emergency conditions, thus improving generalizability of the findings to broader settings and population (5) comprehensive training of all clinical staff in cerebral NIRS monitoring at participating sites, reflecting real-world effective education strategy; (6) including deviations in cerebral oxygenation lasting more than a minute in the analysis which balanced the risk of acting on false alarms from poor sensor contact with the participant and identifying true real-time deviations and (7) a low risk of attrition bias, with only four infants excluded from the analysis after randomization.

This trial had a few limitations. Multiple births beyond twins were excluded due to limited numbers of available NIRS devices required to deliver the intervention. However, no such births occurred at the participating sites, minimizing the practical impact of this exclusion criterion. There is a potential for inadvertent selection bias, as a higher proportion of infants in the intervention group were exposed to chorioamnionitis. These imbalances were addressed through statistical adjustment in the analysis of the primary outcome. Interestingly, despite exposure to chorioamnionitis, often associated with respiratory instability, the intervention improved stability of cerebral oxygenation. The study had no infants who were born <23 weeks’ gestation, therefore the benefit of the intervention in these infants needs exploration.

In conclusion, treatment guided by cerebral NIRS monitoring is a safe, low-risk intervention that significantly improves stability of cerebral oxygenation in extremely preterm infants during the first five days after birth. However, it remains uncertain whether this improvement translates into improved neurological outcomes in early childhood for these infants.^15^ Neurodevelopment outcomes for infants enrolled in this trial are still being collected and they will be reported later. In the SafeBoosC-III trial, treatment guided by cerebral oximetry monitoring in the first 72 hours after birth did not reduce the incidence of mortality or severe brain injury at 36 weeks’ postmenstrual age in extremely preterm infants.^16^ Resolving this uncertainly will require well-conducted RCTs and an individual patient data meta-analysis.^17^ Such trials should incorporate alternate consenting approaches, such as a waiver or deferred consent, in accordance with regional regulatory and ethical compliance.^18^

## Data Availability

All data produced in the present study are available upon reasonable request to the authors, such request is subject to signed data access and authorship agreement with the institution's regulatory body.

## Acknowledgement

We acknowledge funding support from a Project Grant awarded by the Research Foundation, Cerebral Palsy Alliance. We thank Ms Pranali Jani for creating and editing trial education materials used for training staff at the participating sites. We thank all participating families.

## Author contributions

### Concept and design

Pranav R Jani, Traci-Anne Goyen, Himanshu Popat, Rajesh Maheshwari, Kiran Kumar Balegar, Maria Saito-Benz, Tim Schindler, Dharmesh Shah, Melinda Cruz, Manelle Merhi, James Moore.

### Acquisition, analysis, or interpretation of data

Rajesh Maheswari, Dharmesh Shah, Melissa Luig, Daphne D’Cruz, Hayley McDonagh, Mark Tracy, Pranav R Jani, Traci-Anne Goyen, Kiran Kumar Balegar, Maria Saito-Benz, Tim Schindler, James Moore, Aldo Perdomo, Stephanie Morakeas, Vishnu Dasireddy, Mihaela Culcer, Vijay Shingde, Karen Bennington, Joanna Michalowski, Andreja Fucek, Jennifer Querim’, Sean Hanrahan, James Santanelli, James Elhindi, Brian Gloss, Robert Halliday, Himanshu Popat.

### Drafting of the manuscript

Pranav R Jani, Traci-Anne Goyen, Dharmesh Shah, Rajesh Maheshwari, James Elhindi, Himanshu Popat.

### Critical review of the manuscript for important intellectual content

All authors.

### Statistical analysis

James Elhindi, Brian Gloss, Robert Halliday.

### Obtained funding

Pranav R Jani, Traci-Anne Goyen, Himanshu Popat.

### Administrative, technical, or material support

Yang Song, Aldo Perdomo, Stephanie Morakeas.

## Data Sharing Statement

See Supplement.

## References

1. Chow SSW, Creighton, P., Chambers, G.M., Lui, K. Report of the Australian and New Zealand Neonatal Network 2020. Sydney: ANZNN; 2022.

2. Henry G, Webb A, Galea C, et al. Out-of-pocket costs for families and people living with cerebral palsy in Australia. PloS one. 2023;18(7):e0288865.

3. Tonmukayakul U, Shih ST, Bourke-Taylor H, et al. Systematic review of the economic impact of cerebral palsy. Res Dev Disabil. 2018;80:93–101.

4. Achana F, Johnson S, Ni Y, et al. Economic costs and health utility values associated with extremely preterm birth: Evidence from the EPICure2 cohort study. Paediatr Perinat Epidemiol. 2022;36(5):696–705.

5. Yates N, Gunn AJ, Bennet L, Dhillon SK, Davidson JO. Preventing brain injury in the preterm infant—current controversies and potential therapies. Int J Mol Sci. 2021;22(4):1671.

6. Rantakari K, Rinta-Koski O-P, Metsäranta M, et al. Early oxygen levels contribute to brain injury in extremely preterm infants. Pediatr Res. 2021;90(1):131–139.

7. Costa FG, Hakimi N, Van Bel F. Neuroprotection of the perinatal brain by early information of cerebral oxygenation and perfusion patterns. Int J Mol Sci. 2021;22(10):5389.

8. Schmidt B, Whyte RK, editors. Oxygen saturation target ranges and alarm settings in the NICU: what have we learnt from the neonatal oxygenation prospective meta-analysis (NeOProM)? Semin Fetal Neonatal Med. 2020;25(2):101080.

9. Vesoulis ZA, Sharp DP, Lalos N, Swofford DP, Chock VY. Cerebral Near_Infrared Spectroscopy Use in Neonates: Current Perspectives. Res Rep Neonatol. 2024;14:85–95.

10. Hyttel-Sorensen S, Pellicer A, Alderliesten T, et al. Cerebral near infrared spectroscopy oximetry in extremely preterm infants: phase II randomised clinical trial. BMJ. 2015;350:g7635.

11. Jani PR, Goyen TA, Virupakshappa KKB, et al. Targeted Cerebral Oxygenation Using Dedicated Treatment Versus Usual Care in Extremely Preterm Infants: Protocol for a Multicentre International Phase II Randomised Controlled Trial. J Paediatr Child Health. 2025;61(7):1020–1029.

12. Papile L-A, Burstein J, Burstein R, Koffler H. Incidence and evolution of subependymal and intraventricular hemorrhage: a study of infants with birth weights less than 1,500 gm. J Pediatr. 1978;92(4):529–534.

13. Bell MJ, Ternberg JL, Feigin RD, et al. Neonatal necrotizing enterocolitis: therapeutic decisions based upon clinical staging. Ann Surg. 1978;187(1):1–7.

14. Hyttel-Sorensen S, Austin T, van Bel F, et al. A phase II randomized clinical trial on cerebral near-infrared spectroscopy plus a treatment guideline versus treatment as usual for extremely preterm infants during the first three days of life (SafeBoosC): study protocol for a randomized controlled trial. Trials. 2013;14(1):1–8.

15. Hyttel_Sorensen S, Greisen G, Als_Nielsen B, Gluud C. Cerebral near_infrared spectroscopy monitoring for prevention of brain injury in very preterm infants. Cochrane Database Syst Rev. 2017;CD011506 (9).

16. Hansen ML, Pellicer A, Hyttel-Sørensen S, et al. Cerebral oximetry monitoring in extremely preterm infants. N Engl J Med. 2023;388(16):1501–11.

17. Seidler AL, Hunter KE, Cheyne S, Ghersi D, Berlin JA, Askie L. A guide to prospective meta-analysis. BMJ. 2019;367:l5342.

18. De Luca D, Modi N, Davis P, et al. The Lancet Child & Adolescent Health Commission on the future of neonatology. Lancet Child Adolesc Health. 2025;9(8):578–612.

19. Hopewell S, Chan A-W, Collins GS, et al. CONSORT 2025 statement: updated guideline for reporting randomised trials. Lancet. 2025;405(10489):1633–1640.

