## Supplement for "Cerebral Oxygenation Stability In Extremely Preterm Infants: A Randomized Clinical Trial"

eTable 1: Primary outcome using 10 minutes trigger interval stratified by gestational age of the participants

eTable 2: Treatment and complications in participants within the first 5 days after birth

**eTable 1: Primary outcome using 10 minutes trigger interval stratified by gestational age of the participants**

| Outcomes | Intervention group | Standard care group | Relative change in % (95% CI) |
|---|---|---|---|
| **For Gestational ages $23^{+0}$ – $25^{+6}$** | **n = 15** | **n = 14** | |
| Burden of cerebral hypoxia and hyperoxia expressed as % hours, median (interquartile range) | 3.3 (2.3 – 10.9) | 47.2 (34.3 – 91.7) | 170.8 (107.4 to 413.6) |
| Burden of cerebral hypoxia expressed as % hours, median (interquartile range) | 0.0 (0.0 – 0.0) | 1.5 (0.0 – 11.5) | 104.3 (71.3 to 187.4) |
| Burden of cerebral hyperoxia expressed as % hours, median (interquartile range) | 3.3 (2.3 – 10.9) | 47.2 (23.3 – 87.3) | 154.5 (97.8 to 384.5) |
| **Gestational ages $26^{+0}$ – $28^{+6}$** | **n = 35** | **n = 36** | |
| Burden of cerebral hypoxia and hyperoxia expressed as % hours, median (interquartile range) | 4.8 (1.9 – 14.1) | 19.0 (4.9 – 51.3) | 59.0 (42.2 to 100.0) |
| Burden of cerebral hypoxia expressed as % hours, median (interquartile range) | 0.0 (0.0 – 0.0) | 0.0 (0.0 – 1.2) | 37.2 (29.0 to 52.6) |
| Burden of cerebral hyperoxia expressed as % hours, median (interquartile range) | 4.5 (1.9 – 14.1) | 13.4 (4.2 – 44.0) | 45.6 (31.2 to 88.1) |
| **All Gestational ages** | **n = 50** | **n = 50** | |
| Burden of cerebral hypoxia and hyperoxia expressed as % hours, median (interquartile range) | 4.0 (2.0 – 14.4) | 34.5 (6.0 – 76.5) | 93.5 (68.4 to 147.4) |
| Burden of cerebral hypoxia expressed as % hours, median (interquartile range) | 0.0 (0.0 – 0.0) | 0.0 (0.0 – 2.7) | 142.0 (85.8 to 414.3) |
| Burden of cerebral hyperoxia expressed as % hours, median (interquartile range) | 3.5 (2.0 – 14.4) | 23.7 (5.8 – 60.7) | 80.7 (56.8 to 139.1) |

**eTable 2: Treatment and complications in participants within the first 5 days after birth**

| | Cerebral NIRS monitoring + clinical treatment guideline (Intervention group) (n=50) | Blinded cerebral NIRS monitoring + treatment as usual (Standard care group) (n=50) |
|---|---|---|
| Invasive ventilation, No. (%) | 36 (72) | 33 (66) |
| Inhaled nitric oxide, No. (%) | 2 (4) | 2 (4) |
| Pulmonary air leaks, No. (%) | 1 (2) | 1 (2) |
| Postnatal systemic steroids including hydrocortisone, No. (%) | 2 (4) | 5 (10) |
| Treatment for systemic hypotension [a], No. (%) | 11 (22) | 15 (30) |
| Medical treatment for closure of patent ductus arteriosus, No. (%) | 21 (42) | 27 (54) |
| Confirmed sepsis, No. (%) | 4 (8) | 1 (2) |
| Death, No. (%) | 1 (2) | 1 (2) |
| Skin injury at sensor site, No. (%) | 0 | 1 (2) |

Abbreviation: NIRS, near infrared spectroscopy
Data is presented as number (percentage) or median (interquartile range).
[a]Normal saline bolus with or without inotropes
